# Immunophenotype signatures in acute leukemias unveiled by integrative systems immunology

**DOI:** 10.1101/2024.06.19.24309033

**Authors:** Ian Antunes Ferreira Bahia, Rafael Duarte Lima, Gustavo Henrique de Medeiros Oliveira, Antonia Pereira Rosa Neta, Igor Salerno Filgueiras, Lena Schimke Marques, Alexandre Henrique Marques, Leandro M Fonseca Dennyson, Pedro Marçal Barcelos, Adriel Leal Nóbile, Anny Silva Adri, Júlia Nakanishi Usuda, Hans D. Ochs, Haroldo Dutra Dias, Helder I Nakaya, Rodrigo de Souza Barroso, André Ducati Luchessi, Otávio Cabral-Marques, Geraldo Barroso Cavalcanti Junior

**Author notes:** Corresponding authors **Ian Antunes Ferre**i**ra Bah**i**a** Postgraduate Program of Pharmaceuticals Sciences, Federal University of Rio Grande do Norte, Health Sciences Center Avenida General Gustavo Cordeiro Farias, Petrópolis Natal-RN, 59012-570, Brazil. **Geraldo Barroso Cavalcanti Junior** Department of Clinical and Toxicological Analyses, Federal University of Rio Grande do Norte, Health Sciences Center Avenida General Gustavo Cordeiro Farias, Petrópolis Natal-RN, 59012-570, Brazil. **Otavio Cabral-Marques**, MSc, PhD Department of Medicine, Division of Molecular Medicine, University of São Paulo School of Medicine Avenida Dr. Arnaldo, 455 São Paulo, SP, 01246-903, Brazil. Contributed equally (Otávio Cabral-Marques, Geraldo Barroso Cavalcanti Junior).

## Abstract

Acute leukemias (ALs) are complex hematological disorders, and accurate diagnosis is crucial for guiding treatment decisions and predicting patient outcomes. While changes in cell marker levels are well documented, the impact of these changes on marker relationships through an integrative systems approach remains uncharacterized. To address this gap, we conducted a 12-year study investigating 41 markers, including ontogenic markers and those used to diagnose both common and rare leukemia types, using immunophenotyping flow cytometry (IFC) data from 1,069 leukocyte samples obtained from peripheral blood (PB) or bone marrow (BM) aspirates of patients with suspected ALs. Machine learning techniques, such as principal component analysis (PCA) and random forest (RF) classification, demonstrated the stratification power of the cellular markers. Hierarchical clustering analysis of leukocyte ontogenetic markers revealed disease-specific clusters, irrespective of sex or sample type (PB or BM). Additionally, we found that patients with acute myeloid leukemia (AML) showed mild disruption in cell marker correlations, whereas the most significant dysregulation was observed in patients with T-cell acute lymphoblastic leukemia (T-ALL). Importantly, we identified ontogenic correlation changes indicating clusters of immature versus mature leukocyte markers, as well as cell lineage-specific markers influencing cellular relationships. These findings underscore the value of integrating systems strategies into conventional IFC analyses to enhance synthetic diagnosis and deepen our understanding of ALs pathophysiology.

## 1 INTRODUCTION

Acute leukemias (ALs) are characterized by the clonal proliferation of abnormal hematopoietic progenitor cells, exhibiting notable heterogeneity (Weir & Borowitz, 2001). Among the diagnostic tools available, immunophenotyping flow cytometry (IFC) stands out as essential (Weir & Borowitz, 2001). One of the earliest international standards in this field was the French-American-British (FAB) classification, which was based on cytomorphology (Bennett et al., 1976). Building on cytomorphological foundations, IFC employs a panel of monoclonal antibodies (MoAbs) conjugated with various fluorochromes to examine the expression of cluster of differentiation (CD) antigens, allowing for the precise identification of distinct leukocyte populations (Ikoma et al., 2014). The European Group for the Immunological Characterization of Leukemias (EGIL) classification subsequently complemented the FAB system by integrating immunological data from IFC, distinguishing cellular lineages in ALs based on specific markers (Bene et al., 1995).

Recent classifications, including those by the world health organization (WHO), are increasingly incorporating cytogenetic and molecular parameters to classify ALs, reflecting technological advances that enhance diagnostic accuracy (Alaggio et al., 2022a; Barreto et al., 2022; Swerdlow et al., 2016). For instance, in acute promyelocytic leukemia, myeloid antigens may coexist despite the absence of human leukocyte antigens HLA-DR and CD34, along with blast cells exhibiting “cup-like” morphology. The predominant chromosomal translocation t(15;17)(q22;q12) defines the PML-RARA fusion gene mutation in most cases. Additionally, distinctive clinical manifestations, such as hemorrhagic episodes, are expected (Fang et al., 2022; Liu et al., 2013). This exemplifies the complex nature of such AL cases, underscoring the necessity for multidisciplinary and complementary approaches to achieve increasingly precise diagnoses.

To address this gap, systems biology approaches could serve as valuable analytical tools in immunophenotyping, which remains underexplored in this field but is extensively applied in interpreting high-throughput data such as transcriptomes and proteomes. By applying these approaches to IFC data, we can move beyond conventional observation of marker expression and view them as components of interactive immune cell networks across various diagnostic scenarios (Bergthaler & Menche, 2017; Rieckmann et al., 2017). In this context, bioinformatic tools and methods are crucial for facilitating integrative marker analysis, allowing for a holistic interpretation of interdependent components to form a cohesive whole (Beyrend et al., 2018; Bonilha, 2022; Höllt et al., 2016). This approach not only aids in understanding correlations between different marker groups but also enables a more comprehensive data exploration. Moreover, the utilization of machine learning algorithms has further optimized clinical laboratory practice by providing valuable insights and contributing to more precise and efficient decision-making (Lin et al., 2023; Ng et al., 2024; Riva et al., 2023b; Seifert et al., 2023; Zhong et al., 2022).

Drawing from these observations, we employed a stepwise, integrative systems immunology approach to thoroughly characterize the signatures of IFC data from patients with ALs collected over a span of 12 years. This study aimed not only to encompass conventional IFC analysis, which involves observing the expression of each marker within its designated panel, but also to systematically integrate these findings with marker correlations. Recognizing that the immune system operates in an interconnected and synergistic manner (Bergthaler & Menche, 2017; Rieckmann et al., 2017), we utilized a systems immunology approach (**Figure 1**) to provide a more comprehensive understanding of ALs.

**Figure 1.**
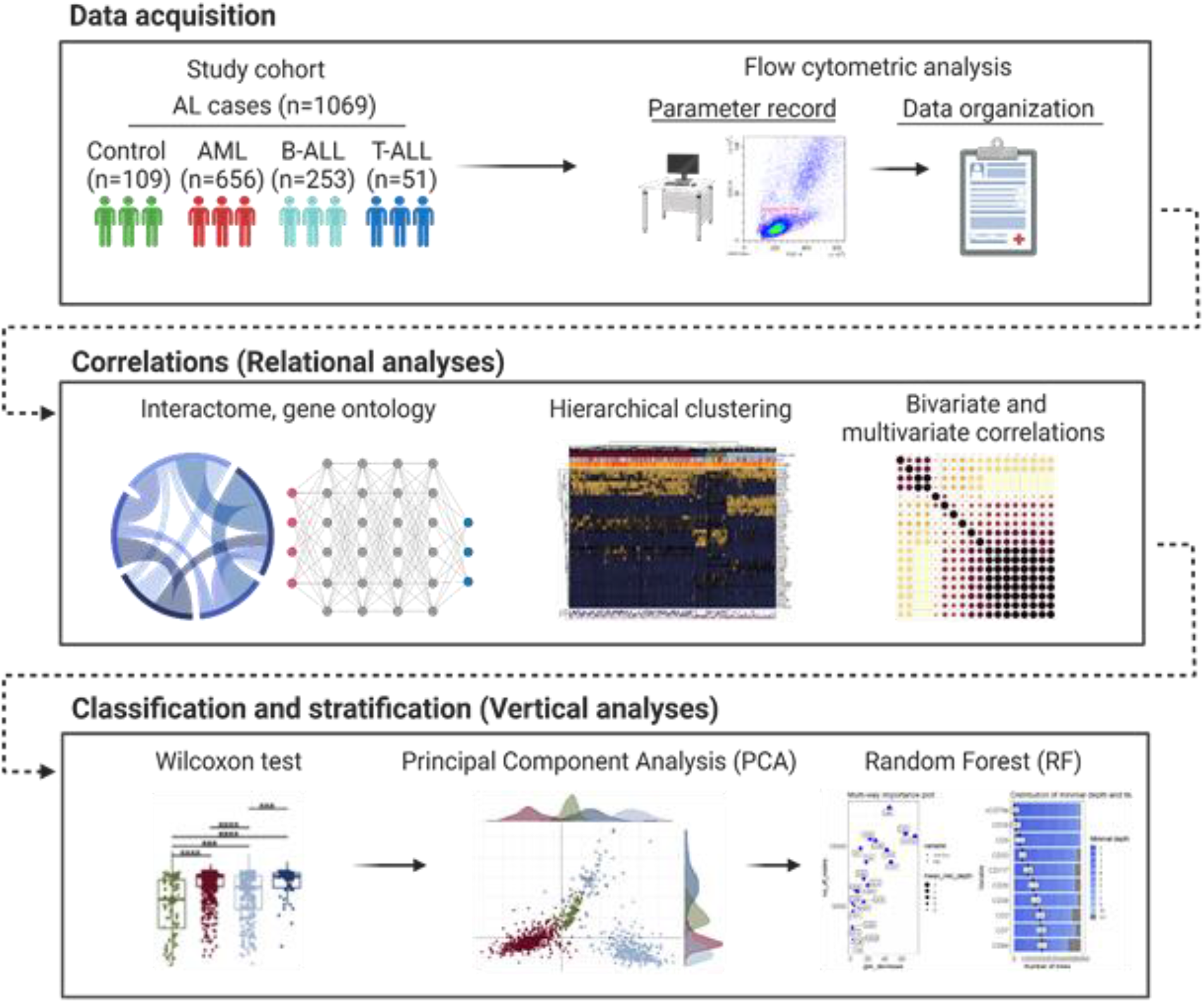

## 2 MATERIALS AND METHODS

### 2.1 Study cohort

A total of 1069 samples (**Supplementary table S0**) of peripheral blood leukocytes (PBL, n = 342) and bone marrow aspirate (BM, n = 727) were collected from patients (442 females and 627 males; mean age = 35) suspected of ALs between 2010 and 2022. Flow cytometric analysis was performed at the Hemocentro Dalton Cunha (HEMONORTE) in Natal/RN, Brazil. Initial diagnoses of ALs were established based on a blast count in the BM exceeding 20%. AL cases were distinguished from other conditions, such as chronic myeloproliferative neoplasms (MPNs) and chronic lymphoproliferative diseases (CLPDs) as previously described (Alaggio et al., 2022a; Khoury et al., 2022). The control group included inconclusive cases, where no pathological or neoplastic populations were detected, though results may vary due to inflammatory phenomena.

This project was approved by the Research Ethics Committee of the Onofre Lopes University Hospital, Federal University of Rio Grande do Norte (CEP/HUOL.UFRN, No 4.378.690). The Hemocenter Dalton Cunha, the institution responsible for the immunophenotypic analyses, granted permission for data use, ensuring informed consent and compliance with ethical and legal standards.

### 2.2 Flow cytometric acquisition

IFC was performed using 4-color panels with leukemia-specific MoAbs as previously described (Ikoma et al., 2014). Our IFC data consisted of 41 flow cytometry markers to phenotypically characterize patients with acute leukemias (B Acute Lymphoid Leukemia [B-ALL], T Acute Lymphoid Leukemia [T-ALL], and Acute Myeloid Leukemia [AML]) as well as our controls. MoAbs were conjugated to fluorochromes including fluorescein isothiocyanate (FITC), phycoerythrin (PE), chlorophyll protein pyridine (PerCP), and allophycocyanin (APC). Laboratory procedures followed the standard protocol provided by the MoAb manufacturers, consistent with established methodologies (Alaggio et al., 2022a; Flores-Montero et al., 2019; Khoury et al., 2022; Lacombe et al., 2016; Matutes, 1995).

Each sample was evaluated using isotype antibodies as negative controls, and an average of 20,000 events per tube were acquired using the FACScan cytometer (Fluorescence Activated Cell Analyzer/FACScan, San Jose, CA, USA). Sample data were analyzed with the Cell Quest software (Becton Dickinson Immunocytometry Systems, San Jose, CA, USA), focusing on mononuclear cells and prioritizing the identification and characterization of blast cell populations. Forward Scatter (FSC-A) and Side Scatter (SSC-A) on a linear scale were considered, along with fluorescence in channels FL1, FL2, FL3, and FL4 on a logarithmic scale. Each parameter’s expression was noted, and the final analysis was confirmed using histograms and dot plots to determine the percentage of positively marked blast cells.

### 2.3 Flow cytometry analysis

We combined two flow cytometry standard (FCS) files from each disease group in our cohort, providing a more representative sample that represent 12 years of immunophenotypic analyses. The collected data underwent rigorous pre-processing and compensation procedures to address marker spillover, utilizing quality control beads and specific cytometer acquisition settings. Subsequently, a standardized logicle transformation was applied to normalize the data (parameters: w = 0.5, t = 1,000,000, m = 4.5).

Implementing a bioinformatics approach, we applied a data processing framework to perform differential analysis of IFC data from our cohort (Hahne et al., 2009; Riva et al., 2023a; Robinson et al., 2023; Seegmiller et al., 2019). The merged FCS files resulted in cellular heterogeneity due to the combined presence of cells from different samples, providing an opportunity to investigate potential variations in cellular distribution across patient groups. After quality control, we employed FlowSOM to identify clusters (Baumgaertner et al., 2021; Piñero et al., 2022; Rasheed et al., 2021; Saeys et al., 2016; Van Gassen et al., 2015). This analysis offered a comprehensive visualization of cell populations distributions across diagnostic groups (AML, B-ALL, T-ALL) compared to controls.

### 2.4 Gene ontology analysis

Gene ontology enrichment analysis was performed on 35 genes associated with the expression of the 41 flow cytometry markers (**Supplementary Table S1**), employing the Enrichr online tool. (Chen et al., 2013; Kuleshov et al., 2016; Xie et al., 2021). The genes corresponding to our 41 markers were identified through research on platforms such as NCBI, using each marker as a keyword to find the relevant genes, as detailed in **Supplementary Table S1**. These genes were categorized based on the predominant characterization of the base markers they represent, including B lymphoid, T lymphoid, myeloid, and common patterns. We assessed these genes for their interactions with related biological processes (BPs) and their expected expression patterns according to the lineage characterized by associated markers (**Supplementary Table S2**). Network analysis was performed using the ggnet package to visualize these interactions. References for all R packages and bioinformatics tools used in this study are listed in **Supplementary Table S3**.

### 2.5 Hierarchical clustering differences in marker expression levels

We generated box plots to display the varying expression levels of 41 flow cytometry markers in acute leukemias (AML, B-ALL and T-ALL) and controls using the R version 4.0.5 (The R Project for Statistical Computing. https://www.r-project.org/), R studio Version 1.4.1106 (R-Studio. https://www.rstudio.com/), and the R packages ggpubr, lemon, and ggplot2. Statistical differences in the marker levels among the groups were evaluated using a two-sided Wilcoxon rank-sum test (Cabral-Marques et al., 2022). Additionally, hierarchical clustering based on euclidean distance complete linkage was performed to examine the expression patterns of the 41 immunophenotypic markers. The results were visualized in a heatmap created with the ComplexHeatmap package, considering variables such as gender, groups, and ages.

### 2.6 Correlation analysis

Correlograms were generated using R packages corrplot and scales, applying Spearman tests to compute correlation coefficients. Boxplots illustrating correlation coefficients were produced using R packages ggpubr, Lemon, and ggplot2 within R Studio. Significance levels were determined via two-sided Wilcoxon rank sum tests and annotated with asterisks (*p ≤ 0.05, **p ≤ 0.01, ***p ≤ 0.001, and ****p ≤ 0.0001). The correlation index for each gene was obtained as follows: correlation Index = {(positive correlation value) − (negative correlation value)}/number of genes. The results were visualized by a hierarchical clustering heatmap based on euclidean distance with complete linkage using the ComplexHeatmap R package as previously described (Cabral-Marques et al., 2022). Additionally, the correlation coefficients of each marker in relation to the others are depicted through box plots.

### 2.7 Principal Component Analysis

We employed principal component analysis (PCA) with spectral decomposition (Lever et al., 2017) to assess the discriminative power of the cell markers, excluding those with missing data, in stratifying our cohort into B-ALL, T-ALL, AML and Control groups. Eigenvalues and eigenvectors exceeding one intercept were considered essential for demonstrating group segregation. PCA was conducted using the scaled expression values of the 22 markers. The PCA analysis was performed with the R packages factoextra (Kassambara A & Mundt F, 2020), ggplot2 (Wickham, 2016) and ggExtra (Attali D & Baker C, 2022).

### 2.8 Random Forest model

We employed random forest model (Liaw & Wiener, 2002) to rank the most relevant markers differentiating among the study groups (AML, B-ALL, T-ALL and control groups). This approach not only aimed to identify the most significant markers for acute leukemia but also explored the potential of employing machine learning in this domain. The random forest model was trained using the features of the R package randomForest, employing 5,000 decision trees. Subsequent analysis assessed variable importance based on criteria such as Gini decrease, number of nodes, and minimum average depth. The model’s performance as a classifier was evaluated using the out-of-bag (OOB) error rate and the Receiver Operating Characteristic (ROC) curve. To validate the model, we divided the dataset into training and testing sets, allocating 2/3 of the observations for training and 1/3 for testing.

## 3 RESULTS

### 3.1 Functional relationships between the flow cytometric markers

Among the 1,069 AL individuals analyzed, 304 were diagnosed with acute ALL, including 253 cases of B-ALL and 51 cases of T-ALL. Additionally, 656 individuals were identified with AML, while the remaining 109 cases comprised our control group, featuring inconclusive diagnoses. Definitive diagnoses were based on clinical criteria, cytomorphological exams, and flow cytometric immunophenotyping as described in the Material and Methods section.

To obtain an integrative overview of the 41 cell markers (**Figure 2a**) and the gene ontology (GO) BPs they are involved in, we performed a GO enrichment analysis. This approach reveals the interconnectedness of the cell markers (**Figure 2b**) and highlighted the diverse cellular lineage functions in which they participate (**Figure 2c**). These functions include T and B cell activation and proliferation, cell development (hematopoiesis and differentiation), and cytokine production (interleukin [IL]-8, IL-2, tumor necrosis factor [TNF]). Notably, during certain stages of the analytical workflow (see **Figures 4b**, **5**, and **6**), 19 of the 41 markers had to be excluded due to a high number of missing data, a characteristic variation observed over the 12 years of analysis. This left 22 focus markers, whose genes and key BPs are highlighted in the circos plot (**Figure 2c**; **Supplementary Tables S4** and **S5**).

**Figure 2.**
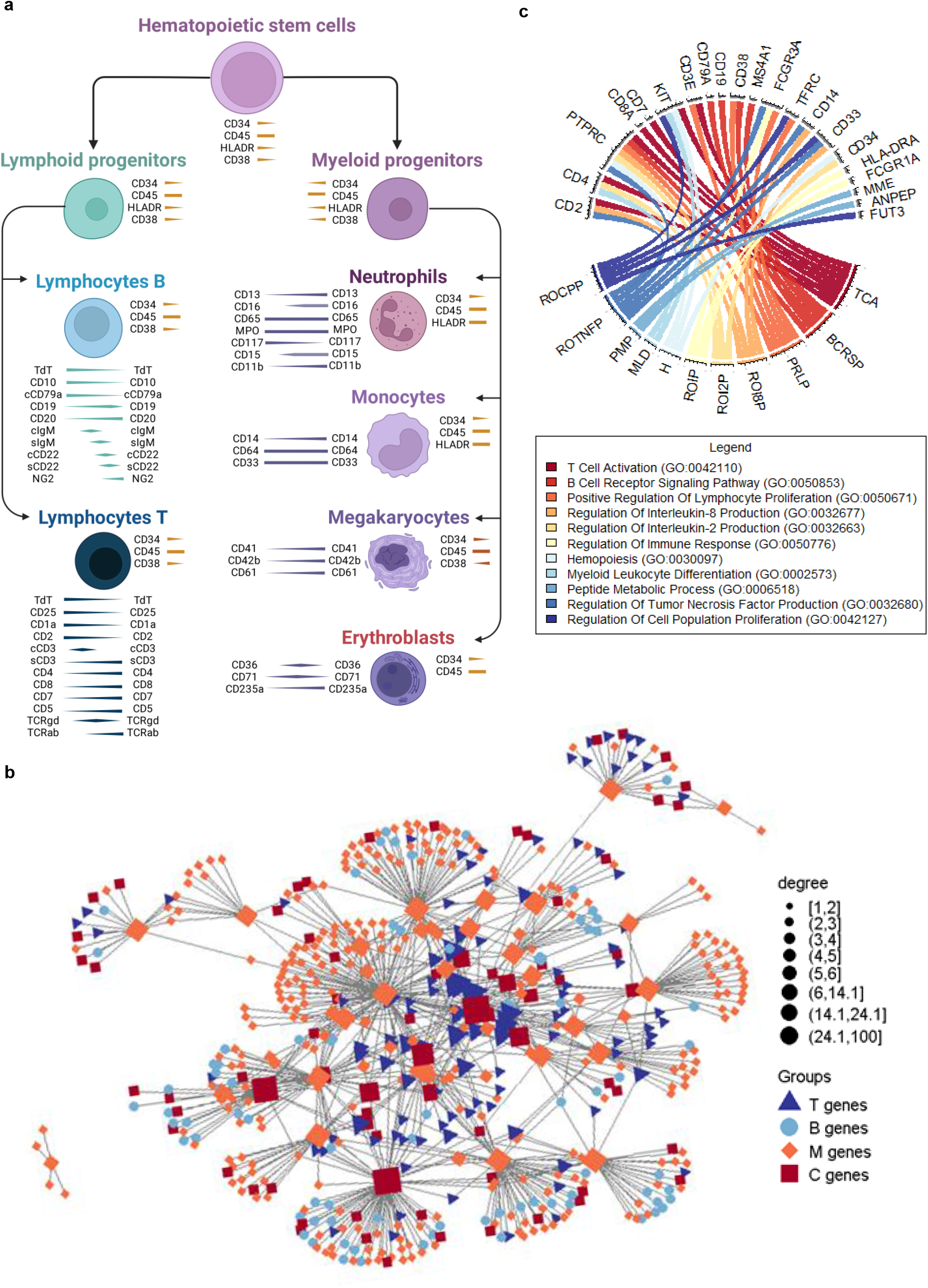

### 3.2 Clustering Acute Leukemia Using Flow Cytometry

To evaluate whether the flow cytometric markers are indeed capable of clustering the ALs groups, we implemented a bioinformatics approach, applying a data processing framework to analyze a set of FCS files. The distribution of cellular data is highlighted in **Figure 3a**, with density plots demonstrating the quality of the readings and the cellular heterogeneity resulting from the file merge according to our cohort. Since the origin of each cell is preserved, this heterogeneity can be further studied post-quality assessment, allowing for the tracking differences in cellular distribution. This process precedes clustering by FlowSOM, a recognized method in this area (Van Gassen et al., 2015). This approach identified 10 metaclusters, enabling comprehensive visualization of the different distributions of cell populations according to the diagnostic groups (AML, B-ALL, T-ALL) in comparison to controls (**Figure 3b-d**).

**Figure 3.**
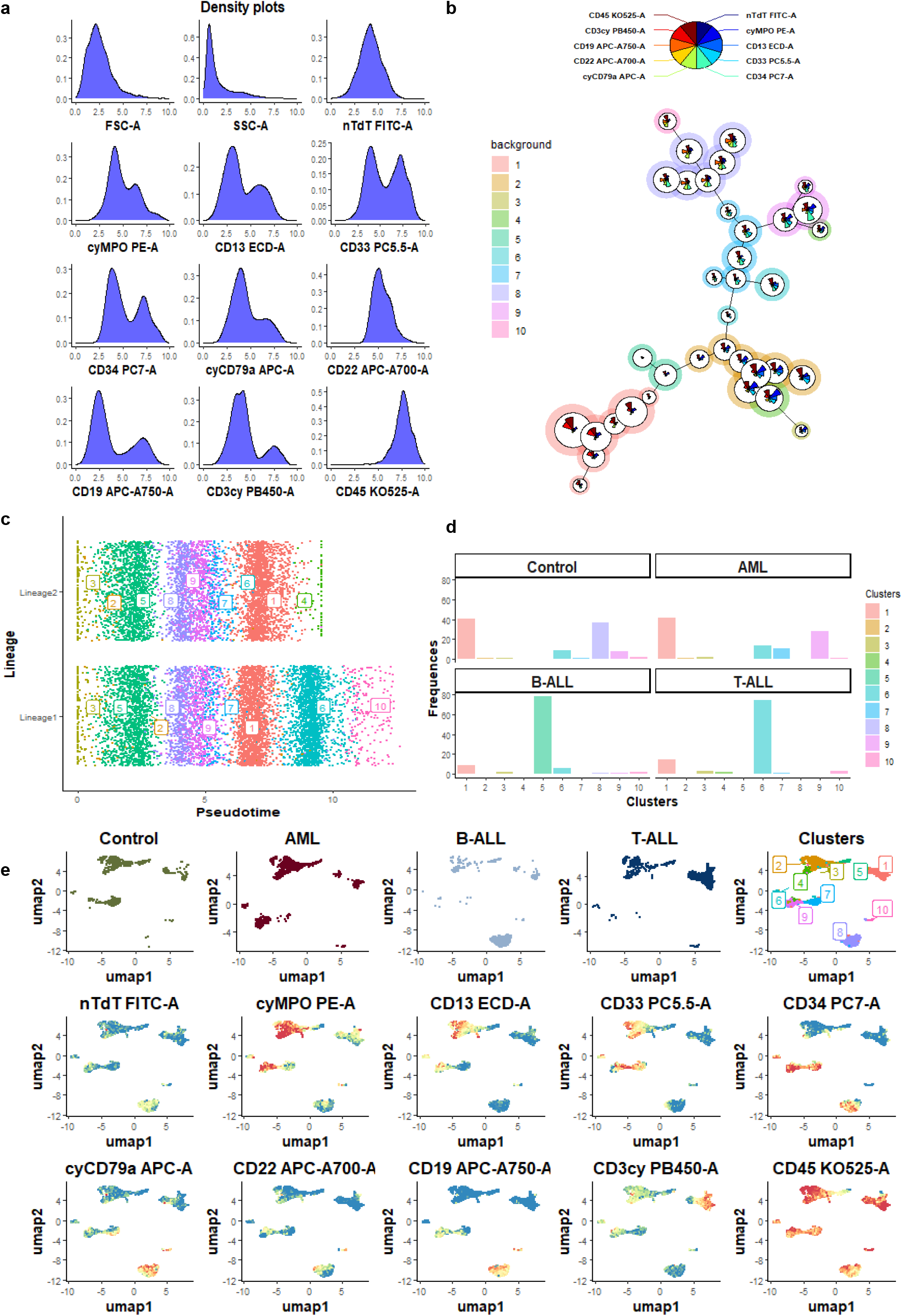

Subsequently, we applied diffusion map analysis with pseudotime calculations to further demonstrate the application of bioinformatics in this domain, providing insights into lineage pattern variations in marker expression intensity (**Figure 3c**). This approach can represent hematopoiesis through IFC. For instance, cluster 9 was predominant in AML, cluster 5 in B-ALL, cluster 6 in T-ALL, and cluster 8 in the control group. Additionally, we identified more generalized distributions between groups, such as cluster 1. These clusters represent characteristic cell populations, such as specific blasts observed in each type of acute leukemia, as well as more general patterns, possibly including neutrophils, which are expected to be present in all cases except those with severely suppressed bone marrow function.

To enhance visualization and cellular identification of clusters, we conducted Uniform Manifold Approximation and Projection (UMAP) analysis, a dimensionality reduction approach (**Figure 3e**). Consistent with the aforementioned analyses, UMAPs enabled us to identify cellular distribution, cluster classification, marker expression density behavior, and cohort variations (ALs vs. control groups).

### 3.3 Distinct immunophenotype and correlation signatures of acute leukemias

To deepen our understanding of the relationships between flow cytometry markers in diagnosing ALs, we conducted hierarchical clustering analysis using lineage-specific markers for the diagnostic groups (AML, B-ALL, T-ALL) compared to controls (**Figure 4a**). This analysis, based on the expression patterns of 41 markers, revealed the significant impact of AL subtypes on the hierarchical clustering signatures (**Figure 4b**). Box plots illustrating the expression distribution of the 41 markers are provided in **Supplementary Figure S1**, whereas **Figure 4c** displays the expression profiles of the 22 markers with the least missing data. Notably, neither patient sex nor sample type (BM or PBL) influenced the clustering patterns of the 41 markers. Furthermore, we observed no *rho* value above 0.4 for all marker when analyzing the relationship between the age and the flow cytometric markers, thus excluding possible age confounding effects **(Supplementary Figure S2 and Supplementary Tables S6-S8)**.

**Figure 4.**
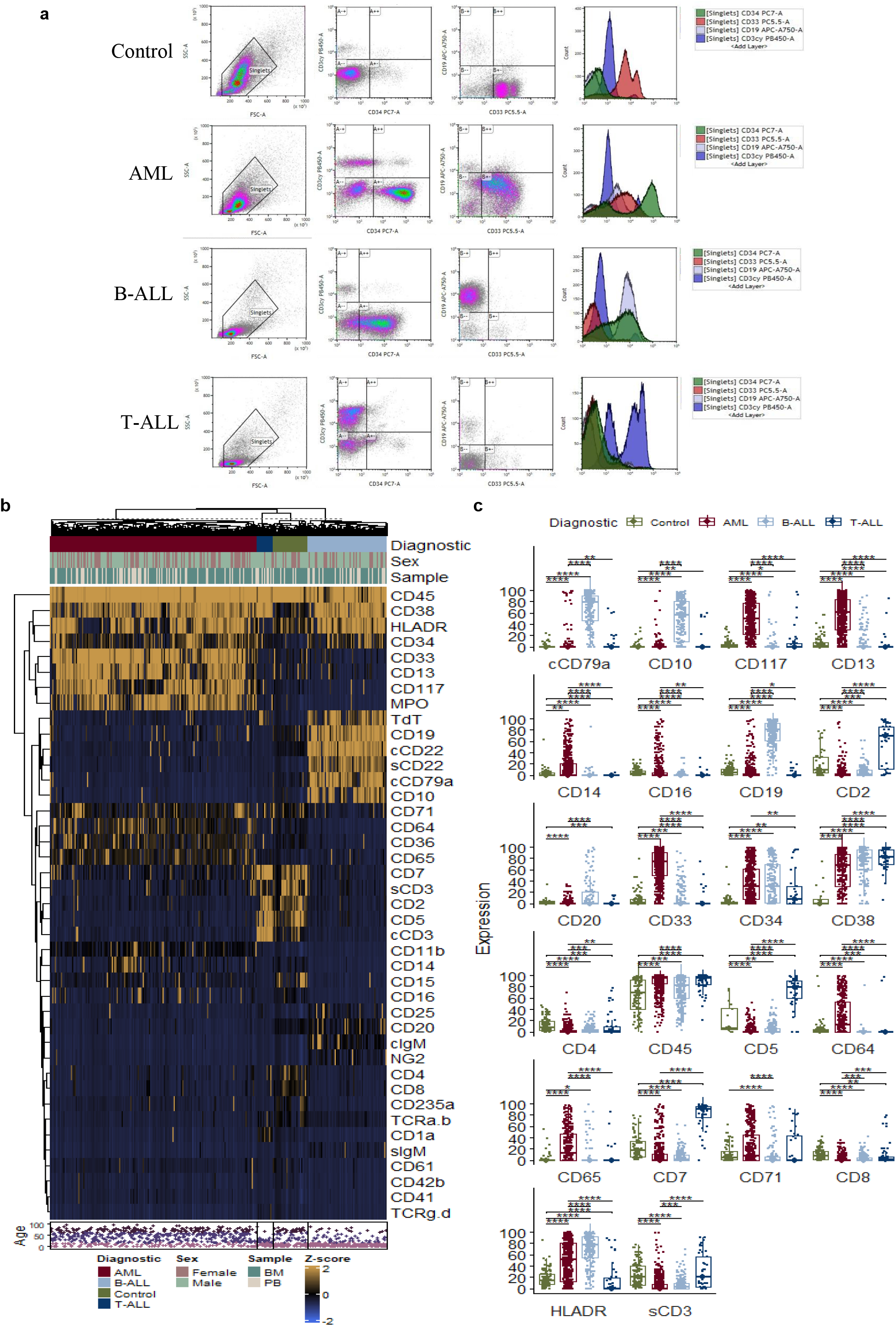

Consistent with the hierarchical clustering analysis, we also observed changes in diagnostic-specific alterations in the correlation signatures of the 22 markers (**Figure 5**; **Supplementary Table S9-S10**). In the AML, there is a significant preservation of relationships between markers of predominantly lymphoid origin (e.g., CD2, sCD3, CD4, CD7, CD8, CD19, and CD20), contrasting with a loss of correlation among myeloid markers (e.g., CD13, CD14, CD16, CD33, CD64, CD65, CD71, and CD117) markers (**Figure 5b**).

**Figure 5.**
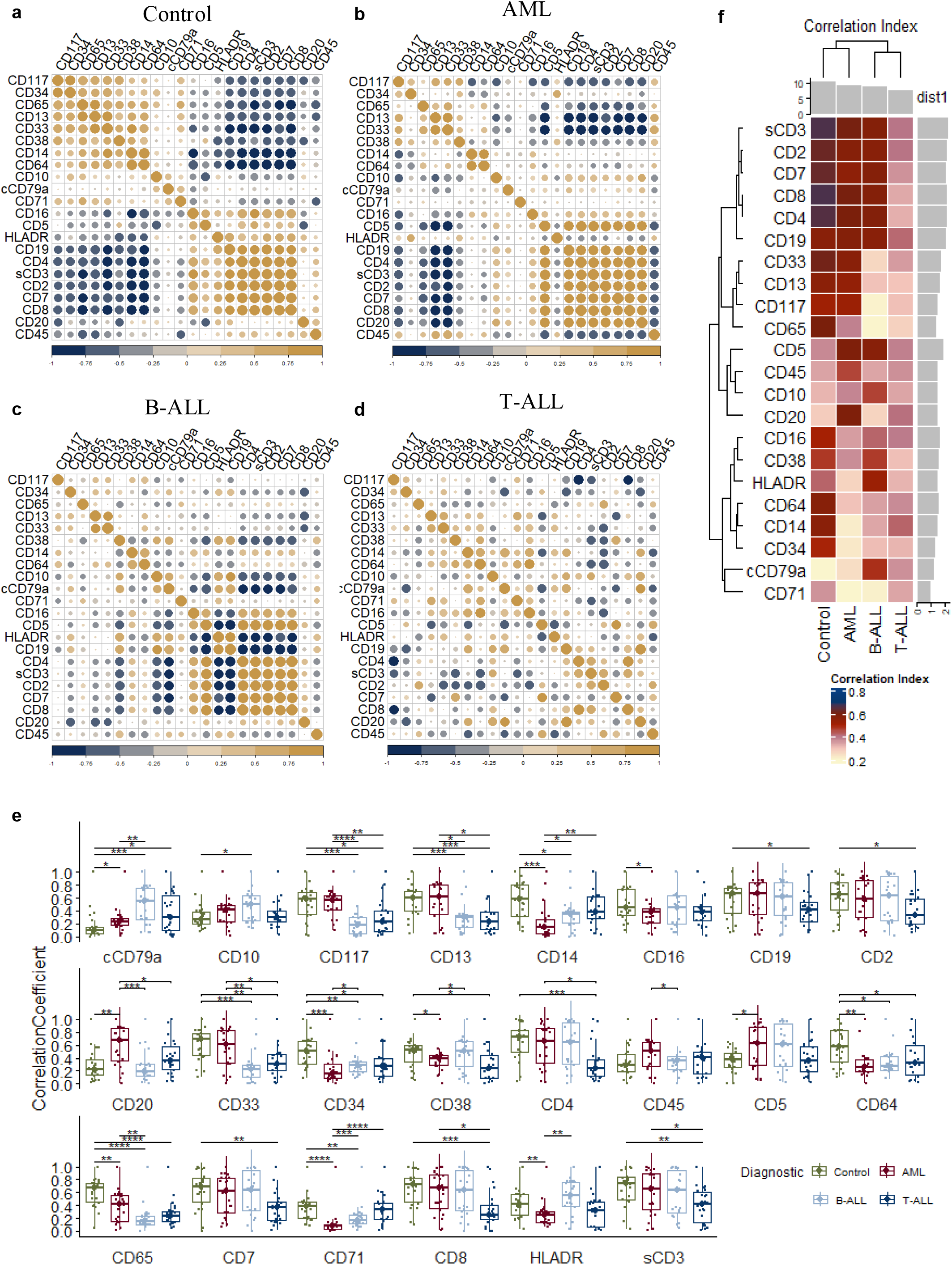

In the B-ALL group, the relationships among the markers typical of T lymphocytes are maintained, while disruptions are evident for markers characteristic of B lymphocytes (e.g., CD19 and CD20) and myeloid cells (**Figure 5c**). On the other hand, the T-ALL group exhibits an almost complete breakdown of relationships among markers, signaling a notable and deeper systemic dissociation compared to the other groups (**Figure 5d**). **Figures 5e** and **5f** display the correlation coefficients and indices of each marker according to the groups.

### 3.4 Ranking flow cytometry markers as predictors of ALs

We assessed the ability of the 22 markers with the least missing data across groups to differentiate between AL patients and controls, while also evaluating the correlations between the flow cytometry markers. To address this, we employed PCA analysis using the spectral decomposition approach (Lever et al., 2017). This analysis demonstrated the discriminatory power of the markers in distinguishing between AML, B-ALL, T-ALL, and control groups (**Figure 6a**). The eigenvalues and marker contributions to different PCA dimensions are detailed in **Supplementary Tables S11** and **S12**, respectively. The variable contribution graph clearly illustrates that myeloid cell markers (CD14, CD16, CD33, CD64, and CD117), B cell markers (CD20, CD19, and cCD79a), and T cell markers (CD2, sCD3, CD4, CD5, CD7, and CD8) effectively stratify AML, B-ALL, and T-ALL into distinct dimensional trajectories from the controls.

**Figure 6.**
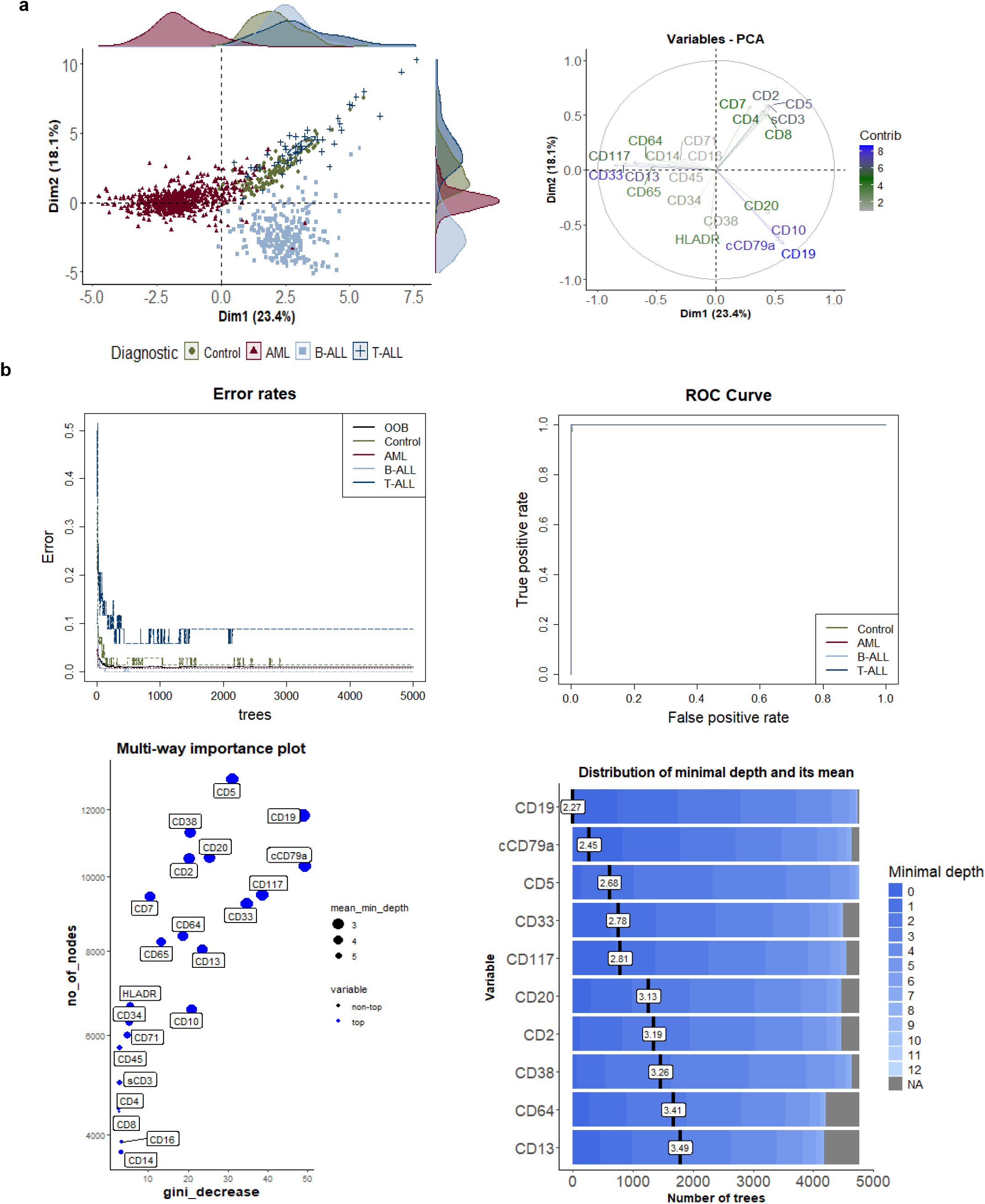

To identify the best predictive markers for the AL groups, we employed a random forest model. The 22 markers were ranked based on their importance in distinguishing ALs from the control group. Details of the model’s confusion matrix, as well as the testing and training groups, are provided in **Supplementary Tables S13** and **S14**, respectively. Consistent with the PCA results, the RF analysis showed an out-of-bag (OOB) error rate of 1.7% and an area under the curve (AUC) of 0.999 for the receiver operating characteristic (ROC) curves when comparing ALs to the control group. These results indicate the high accuracy of the random forest analysis. The distribution of the top 10 markers (**Figure 6b**) identified B cell markers (CD19 and cCD79a), T cell markers (CD5), and myeloid markers (CD33) as having the highest scores for classifying the AL groups.

## 4 DISCUSSION

This study presents a novel approach to the diagnosis of ALs by integrating systems immunology (Cabral-Marques et al., 2022; Prado et al., 2023; Salgado et al., 2021; Schimke et al., 2022) with IFC data. This innovative method not only examines individual marker expressions but also explores the relationships between these markers, providing a more comprehensive understanding of the immune system’s alterations in ALs. The hierarchical clustering analysis revealed distinct immunophenotypic signatures for different types of ALs, which were consistent across sex and sample origin. This finding agrees with previous researches that has shown the robustness of immunophenotypic markers in distinguishing between AL subtypes (Bene et al., 1995; Ikoma et al., 2014; Lacombe et al., 2016). The observation that AML patients exhibited a milder disruption in cell marker correlations compared to T-ALL patients is an interesting finding that may reflect the differential impact of the leukemic process on the immune system. Additionally, the study’s identification of changes in ontogenic correlation patterns is consistent with the concept that ALs are diseases of cell differentiation and maturation (Alaggio et al., 2022b; Khoury et al., 2022). The application of machine learning algorithms, such as PCA and RF classification, to a large dataset of 1,069 samples over a 12-year period underscores the potential of these advanced analytical tools in enhancing diagnostic accuracy. This integrative systems approach not only improves the diagnostic precision of IFC markers but also enriches our understanding of the immunological intricacies and systemic interactions in ALs, potentially offering a valuable addition to the AL diagnosis.

Enrichment analysis of the IFC markers revealed interconnected BPs related to the expression of the 35 genes associated with the markers under study, underscoring their involvement in AL pathogenesis. These processes encompass key aspects of the immune system, including T and B cell activation and proliferation, which are crucial for mounting an effective immune response against pathogens and malignancies (Chapman & Chi, 2022; Chi et al., 2024; Horii & Matsushita, 2021). Additionally, the analysis highlighted relationships for these markers in cell development (hematopoiesis and differentiation), processes frequently disrupted in leukemia (Ahmad et al., 2023; Zhao et al., 2023). Furthermore, cytokine production, including IL-8, IL-2, and TNF, also emerged as a prominent function associated with the analyzed markers. Cytokines play a complex role in the immune system, influencing cell growth, differentiation, and activation (Cao & Kagan, 2023; Cui et al., 2024; Dantzer, 2004; Kelley et al., 2003; Miller et al., 2009; Saxton et al., 2023; Szelényi, 2001). These findings provide a valuable foundation for future research aiming to understanding how disruptions in the interplay between these markers might contribute to the development and progression of ALs.

To further explore cellular differentiation patterns and lineage relationships, the study incorporated diffusion map analysis with pseudotime calculations (Haghverdi et al., 2015). This approach modeled the progressive changes in marker expression intensity that occur during cell differentiation, providing valuable insights into hematopoietic trajectories within the context of ALs. For instance, we identified specific clusters associated with each leukemia type (cluster 9 for AML, cluster 5 for B-ALL, and cluster 6 for T-ALL) alongside more generalized distributions present across all groups (cluster 1). These clusters likely represent characteristic cell populations, such as blasts specific to each leukemia subtype, as well as more common cell types like neutrophils, which are typically present unless bone marrow function is severely compromised (Ahmad et al., 2023).

Dimensionality reduction analysis was employed to enhance visualization and cellular identification accordingly the clusters (Becht et al., n.d.; McInnes et al., 2018). Consistent with previous analyses, we were able to effectively capture cell distribution patterns, cluster classification, variations in marker expression intensity, and differences between patient groups in our cohort. These findings emphasize the power of bioinformatics tools in analyzing flow cytometry data to differentiate between distinct AL subtypes and controls (Beyrend et al., 2018; Cheung et al., 2022; Çubukçu et al., 2023; Melsen et al., 2020; Montante & Brinkman, 2019; Saeys et al., 2016). The ability to identify leukemia-specific cellular populations and visualize differentiation trajectories can significantly optimize the diagnostic process (Ng et al., 2024; Nguyen et al., 2023; Seifert et al., 2023; Simonson et al., 2022).

Delving deeper into the immunophenotype and correlation signatures of cellular populations in IFC, we employed hierarchical clustering analysis (Botta et al., 2022; Prado et al., 2023). Understanding how marker expression patterns and correlations differ between healthy and leukemic cells can provide valuable insights into disease biology and potentially guide the development of novel diagnostic strategies. Our analysis revealed significant impacts of AL clustering patterns, highlighting the distinct immunophenotypes associated with each leukemia type. Notably, neither patient sex nor sample type (BM or PBL) significantly influenced the clustering patterns. Furthermore, an analysis to assess the potential confounding effect of age revealed no significant correlation between age and marker expression (rho value < 0.4 for all markers). Thus, indicating that these parameters do not qualify as confounding variables affecting the observed immunophenotype differences of ALs (Zhou, 2021).

Building upon the clustering analysis, the study investigated how correlations between these markers differ across diagnostic groups, focusing on the 22 markers with minimal missing data. Analyzing marker correlation patterns generates deeper insights into the mechanisms underlying AL development. The observed disruptions in correlations provide strong evidence for dysregulated cellular communication within leukemic blasts (Bewersdorf & Zeidan, 2020; Masoumipour et al., 2021). Our findings revealed intriguing patterns of disrupted correlations within specific lineages for each AL group. In the AML group, we observed notable preservation of correlations among markers predominantly expressed by lymphoid cells and disruption between myeloid markers. This suggests a potential compartmentalization within the leukemic process of AML, where the malignant blasts primarily affect the myeloid lineage while leaving the lymphoid lineage relatively intact (Jaddaoui et al., 2022; Khoury et al., 2022; Meena et al., 2022). The B-ALL group displayed a different pattern, with maintained correlations between T lymphocyte markers but disruptions observed for B lymphocyte and myeloid markers. This finding reflects a more lineage-specific disruption in B-ALL, indicating an impact on B-cell development during leukemogenesis in this AL (Seegmiller et al., 2019). The most significant observation is in the T-ALL group, where an almost complete breakdown of correlations across all markers is evident. This near-complete loss of correlation suggests a more profound and systemic disruption of the immune landscape in T-ALL compared to the other subtypes. These findings on disrupted marker correlations further support the concept of ALs as diseases of aberrant differentiation and maturation (Colom Díaz et al., 2023; Sell, 2005). The differential patterns of disruption observed across subtypes potentially reflect the unique underlying biology of each leukemia type (Harris et al., 2019; Juliusson & Hough, 2016; Weijie, 2022). Future studies can leverage these findings to explore disrupted correlations and their functional consequences, potentially leading to new understandings within AL subtypes.

To assess the discriminatory power of these markers, we employed PCA with spectral decomposition (Costa et al., 2010; Lever et al., 2017; Zhou, 2021). This analysis effectively separated the AL groups (AML, B-ALL, T-ALL) from controls in a multidimensional space. This result further solidifies the concept of distinct immunophenotypes for each AL subtype (Fang et al., 2022; Matutes, 1995; Piñero et al., 2022; Sonneveld et al., 2003). An RF model was also employed to rank the 22 markers based on their effectiveness in differentiating ALs from controls. The model achieved an impressive OOB error rate of 1.7% and an area under the curve (AUC) of 0.999 for the ROC curve, indicating exceptional accuracy. The analysis also highlighted the top 10 most important markers, which included B cell markers (CD19 and cCD79a), a T cell marker (CD5), and a myeloid marker (CD33) (Ikoma et al., 2014; Lacombe et al., 2016). These findings reinforce the importance of a well-established panel for accurate AL diagnosis, paving the way for further investigation into the development of IFC-based marker strategies, as well as the construction and validation of analysis panels in IFC (Alaggio et al., 2022b; Flores-Montero et al., 2019; Khoury et al., 2022).

## 5 STUDY LIMITATIONS

However, it is important to acknowledge several limitations of our study. Firstly, missing data necessitated the exclusion of some markers, which underscores the critical need for standardized data acquisition and management protocols in future research. This would help ensure the completeness and reliability of datasets, thereby enhancing the robustness of the findings. Additionally, our focus on specific markers, while necessary for targeted analysis, may have inadvertently restricted the comprehensiveness of our study. A broader, more inclusive approach could potentially uncover additional insights and interactions that were not captured in our current analysis. This limitation highlights the importance of employing diverse and comprehensive panels of markers in future investigations to obtain a more holistic view.

Moreover, while our findings are promising, they require further validation through prospective studies and experimental confirmation. This step is crucial to establish the robustness and clinical applicability of our results. Experimental validation can provide a deeper understanding of the biological relevance of the identified markers and their functional roles in the context of the studied condition. While acknowledging these limitations, we emphasize the importance of continued research efforts and the application of standardized methodologies to overcome these challenges. Such endeavors will be instrumental in validating and expanding upon our current findings, ensuring their relevance and applicability in clinical settings.

## 6 CONCLUSION

This study presents an innovative diagnostic approach for acute leukemias (ALs) by integrating systems immunology with immunophenotypic data from IFC. Through the analysis of both individual marker expressions and their interrelationships, distinct immunophenotypic signatures for AL subtypes are identified, reinforcing the robustness of these markers. Advanced machine learning techniques reveal the potential for improved diagnostic accuracy. Despite limitations like missing data and marker specificity, the study provides a solid framework for future research. Overall, this integrative approach enhances diagnostic precision and deepens our understanding of the immune system’s role in ALs, offering prospects for the development of novel diagnostic and therapeutic strategies.

## Data Availability

All data used in this study are provided in the Supplementary Data. Raw data (FCS files) are available upon reasonable request.

## Author Contributions

IAFB and OCM wrote the manuscript; IAFB, OCM, GBCJ, DLMF, GHMO, HDD, HIN, ADL, ISF and RDL provided scientific insights; IAFB, OCM, DLMF, AHM, ASA, PMB, ALN, JNU and APRN performed data and bioinformatics analyses; HDD, HIN, ADL, ISF, LSM and HDO revised and edited the manuscript; OCM and GBCJ supervised the project.

## Supporting information

Figures Titles and Legends

## Acknowledgments

We thank the Coordination of Superior Level Staff Improvement under Academic Excellence Program (CAPES/PROEX grants 88887.954341/2024-00 to IAFB, 88887.917898/2023-00 to JNU and 88887.801068/2023-00 to ALN). We thank the National Council for Scientific and Technological Development (CNPq) Brazil (grants: 309482/2022-4 to OCM). We acknowledge the São Paulo Research Foundation (FAPESP grants 2018/18886-9, 2020/01688-0, and 2020/07069-0 to OCM, 2020/16246-2 and 2023/06086-6 to PMB, 2023/07806-2 to ISF, 2020/16246-2 and 2023/13356-0 to DLMF, 2023/12268-0 to ASA, 2023/14417-2 to JNU and 2018/14933-2 to HIN) for financial support. We recognize and thank the infrastructure support and raw data for the Hematology Center Dalton Cunha and Flowmentor.

---

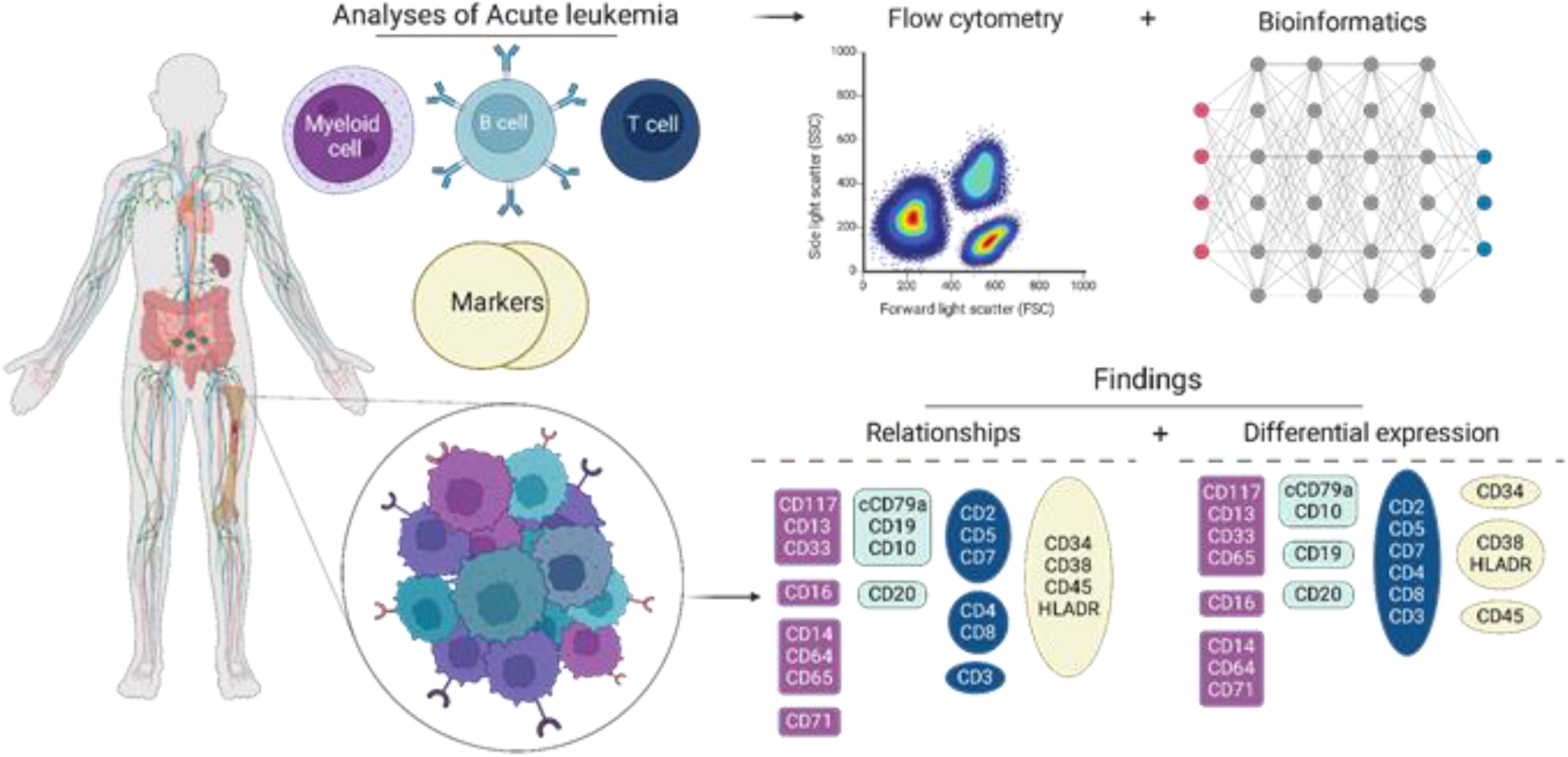
Graphical abstract

**Supplementary figure 1.**
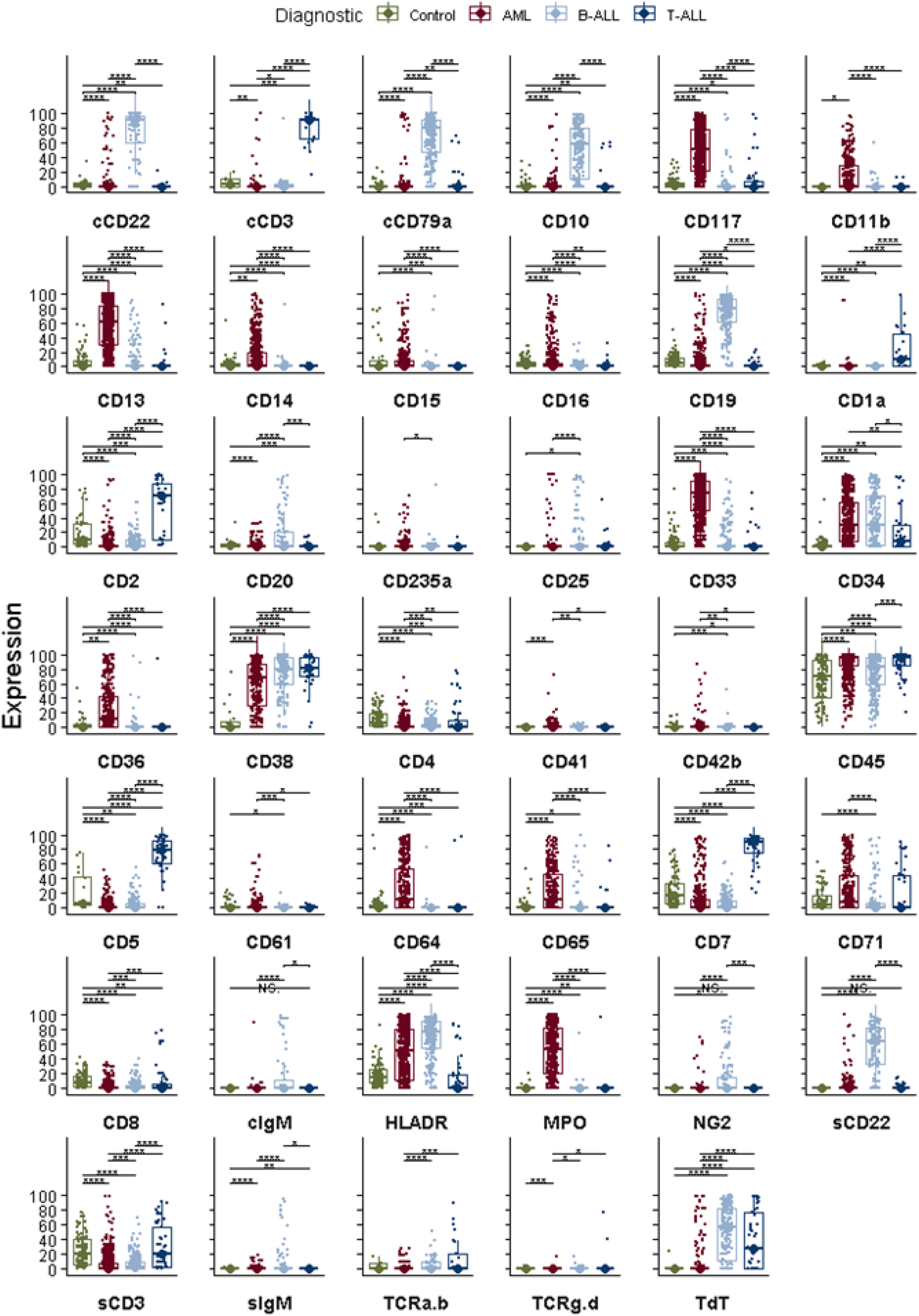

**Supplementary figure 2.**
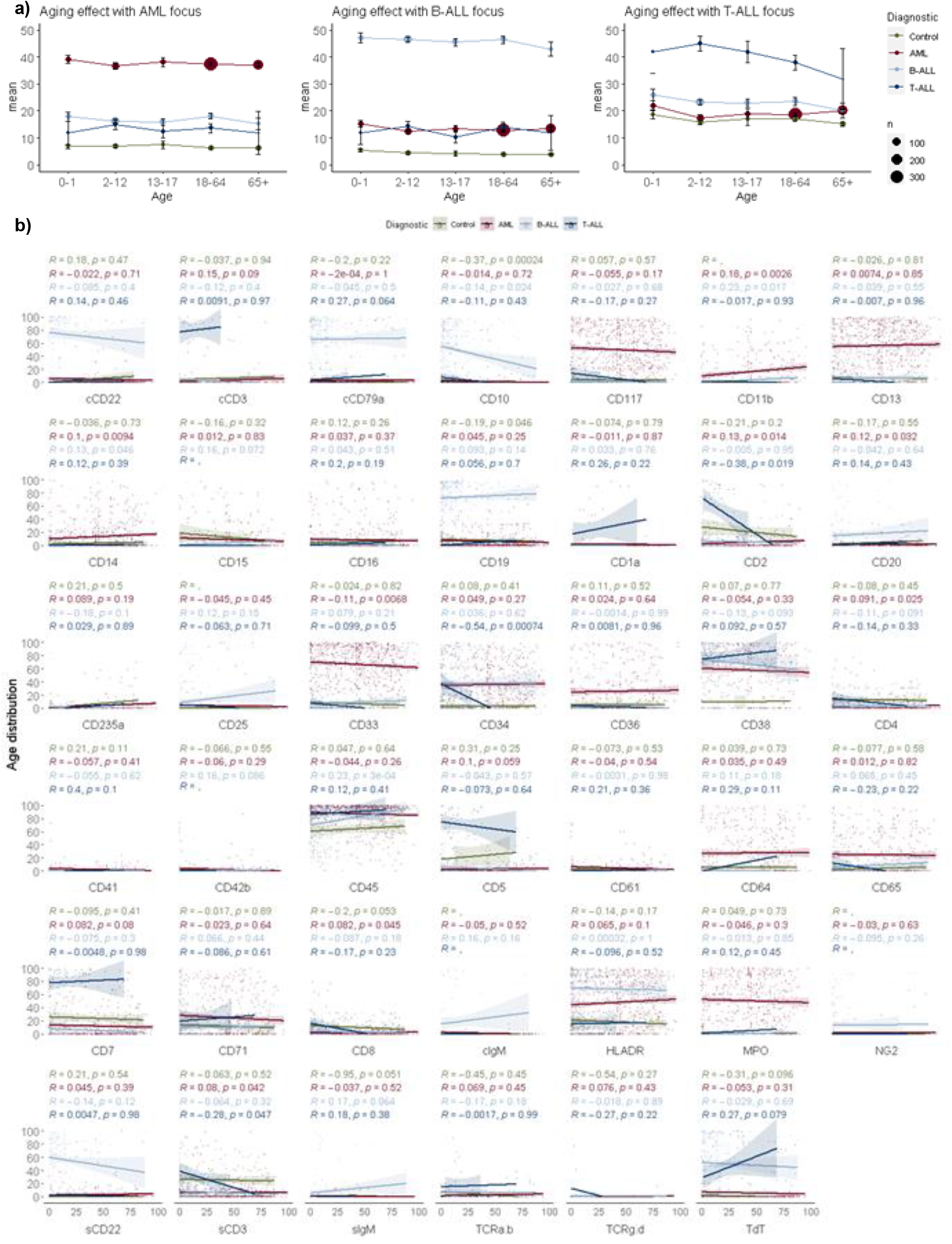

