## Supplementary material for "Immunophenotype signatures in acute leukemias unveiled by integrative systems immunology": Figures Titles and Legends

**FIGURE LEGENDS**

**Main Figures**

**Figure 1. Study workflow.** Following data acquisition (see data and descriptive statistics in **Supplementary Tables S0 and S6**), several bioinformatic methods were applied for the relational (correlations) and vertical (classification and stratification) analyses. (Bioinformatic tools and R packages in **Supplementary Table S3**). Created with BioRender.com.

**Figure 2. Immunophenotyping Marker Landscape, Ontogenetic Patterns and relationships. a)** The image portrays the 41 cell markers used for immunophenotyping of patients with AL, encompassing ontogenic molecules and commonly used diagnostic markers (e.g., common markers for ALs: HLA-DR, CD34, CD38, CD45, TdT; lineage-specific markers: CD1a, CD3, CD2, CD5, CD7, CD4, CD8, CD10, CD19, CD79a, CD20, CD13, CD16, CD33, CD65, MPO, CD117, CD15, CD14, CD64 ), as well as rarely used markers for some specific leukemia types (e.g., αβ and γδ TCRs, CD22, IgM, NG2, CD71, CD36, CD235α, CD41, CD42b, CD61, CD25, CD11b). **b)** The interaction network illustrates the relationship between enriched Gene Ontology (GO) biological process (BP) terms and genes associated with the 41 cellular markers identified using Enrichr. The network comprises a total of 674 BPs. Genes were grouped according to the cell type marker pattern characterized as follows: B lymphoid (B), T lymphoid (T), myeloid (M), or common expression (C) patterns as denoted by different colors and symbols. The size of each symbol represents the degree of relationship. **c)** Circos plot illustrating interactions between 22 genes encoding the main markers with lower heterogeneity among groups (upper half of the circos plot) and primary immune pathways (lower half of the circos plot) denoted by abbreviations and different colors according to figure legend. (Details for this figure is provided in **Supplementary Tables S0-S2, S4 and S5**).

**Figure 3. Comprehensive Analysis of Flow Cytometry Data: From Marker Density Plots to Clustering and Pseudotime Analysis. a)** Density plots representing flow cytometry analysis of IFC markers after data compensation and common logicle transformation (with parameters w = 0.5, t = 1,000,000, and m = 4.5) for some representative CD markers. **b)** FlomSOM clustering with 10 metaclusters selection and fluorescent markers. **c)** Lineage variation represented through pseudotime calculations based on diffusion mapping characterizing patterns of cell markers intensity variations. The analysis is also anchored by clusters. **d)** Heterogeneity study of 10 metacluster data distribution according to our cohort. These clusters represent characteristic cell populations, including subtype-specific blasts and common cell types such as neutrophils. **e)** UMAP application signaling cell distribution according to cluster classification, differential variations across our cohort, and marker expression density.

**Figure 4. Flow Cytometric Marker Expression, Diagnostic Group Variability, and Age-Related Patterns of Acute Leukemia Populations. a)** Representative flow cytometry analysis gating on different cell markers and cell types, delineating acute leukemia populations primarily within mononuclear cells, while covering other subpopulations. Additionally, the percentage expression associated with each cellular marker was shown, adapted to the context of each case and panel. One representative sample for each group is shown. **b)** The heatmap illustrates the expression patterns of the 41 immunophenotypic markers across four diagnostic groups. The variables sex, sample type, and diagnostic group are shown in different colors according to the figure legend in the bar graph above the heatmap. Hierarchical clustering was applied to the different diagnostic groups. The age distribution of individuals in each diagnostic group is outlined in the dot plot below the heatmap (detailed age-related expression patterns are shown in **Supplementary Tables S6**, **S7,** and **S8**). **c)** The boxplots depict expression patterns of the top 22 immunophenotypic markers within the studied groups (**Supplementary Table S15**). Statistical significance between groups was assessed using Wilcox tests and annotated with asterisks (*p ≤ 0.05, **p ≤ 0.01, ***p ≤ 0.001, and ****p ≤ 0.0001) contributing to delineating expression patterns.

**Figure 5. Interrelationship Analysis of Marker Expression Patterns in Different Diagnostic Groups of Leukemia. a-d)** The correlation matrix depicts the interrelationship among the 22 markers exhibiting expression heterogeneity (**Supplementary Table S15**) in each diagnostic group: **a)** Control group, **b)** Acute Myeloid Leukemia, **c)** Acute B Lymphoid Leukemia, and **d)** Acute T Lymphocytic Leukemia (Details in **Supplementary Table S9**). Values ranging from -1 to 1 express the Spearman correlation coefficient, representing the strength and direction of correlation. **e)** Boxplots showing each marker's expression value across the different diagnostic groups. Statistical significance between groups was assessed using Wilcox tests and annotated with asterisks (*p ≤ 0.05, **p ≤ 0.01, ***p ≤ 0.001, and ****p ≤ 0.0001). **f)** A consolidated correlation matrix combining all correlation values from **a-d)** for the respective markers and four diagnostic groups. Correlation values span from 0 to 1, with higher values denoting positive correlations between markers (**Supplementary Table S10**). These visualizations provide individual insights into marker expression patterns, aiding in understanding their relationships within and across diagnostic groups.

**Figure 6. PCA Segregation and Random Forest Classification Contributing to Leukemia Diagnosis. a)** PCA results, showing the contribution of the 22 markers to segregating the 1.069 samples based on the four diagnoses. Detailed eigenvalues for the 22 dimensions are provided in **Supplementary Table S11**, while the individual contributions of each marker are provided in **Supplementary Table S12**. **b)** Construction and results of the Random Forest machine learning model showing the error rate and ROC curve for characterization, categorizing the 22 markers based on their differential expression in the four groups analyzed. The model's essential results are encapsulated in **Supplementary Table S13**, with a detailed representation of the confusion matrix for training and testing samples in **Supplementary Table S14**. Graphs below show the multi-way importance and distribution of minimal depths plot obtained by random forest classification analysis based on the Gini decrease and number of trees for each variable (22 markers with the least heterogeneity; **Supplementary Table S15**), respectively. The top of the graph highlights the top variables differentiating the patient group compared to the control group.

**Supplementary Figures**

**Supplementary Figure 1. Comprehensive Analysis of Immunophenotypic Marker Expression Across Diagnostic Cohorts.** Expression patterns of all 41 immunophenotypic markers within the studied groups are shown, offering a comprehensive view of marker distributions and variations across different diagnostic cohorts.

**Supplementary Figure 2. Age-Dependent Variation in flow cytometric Marker Expression Across Leukemia.** **a)** Distribution of CD marker expression from of patients grouped by age across the four cohorts, demonstrating variations in the average expression levels of key markers characteristic of each leukemia type within five age ranges (in details described in **Supplementary Table S7**. **b)** Linear regression analysis for each marker considering age distribution in each diagnostic groups. Further statistical details provided in **Supplementary Table S8**.
